# The Impact of COVID-19 Pandemic on Cardiology Services

**DOI:** 10.1101/2020.06.10.20126458

**Authors:** Omar Fersia, Sue Bryant, Rachael Nicholson, Karen McMeeken, Carolyn Brown, Brenda Donaldson, Aaron Jardine, Valerie Grierson, Vanessa Whalen, Alistair Mackay

**Affiliations:** Cardiology Department, Dumfries and Galloway Royal Infirmary, UK; Service Improvement, Acute and Diagnostics Department, Dumfries and Galloway Royal Infirmary, UK; Clinical Imaging Department, Dumfries and Galloway Royal Infirmary, UK

## Abstract

**Objective:** COVID-19 pandemic resulted in prioritisation of NHS resources to cope with the surge in infected patients. However, there have been no studies in the UK looking at the effect of COVID-19 work pattern on the provision of cardiology services. We aim to assess the impact of the pandemic on the cardiology services and clinical activity.

**Methods:** We analysed key performance indicators in cardiology services in a single centre in the UK in the intervals prior and during the lockdown to assess for reduction or changes to service provisions.

**Results:** There has been more than 50% drop in patients presenting to cardiology and those diagnosed with myocardial infarction. All cardiology service provisions sustained significant reductions which included outpatient clinics, investigations, procedures and cardiology community services such as heart failure and cardiac rehabilitation.

**Conclusion:** As ischaemic heart disease continues to be the leading cause of death nationally and globally, cardiology services need to prepare for a significant increase in workload in the recovery phase and develop new pathways to urgently help those adversely affected by the changes in service provisions.

**Key Questions:**

- **What is already known about this subject?** COVID-19 affected the way healthcare is delivered through restructuring and prioritisation of resources. It is therefore expected that COVID-19 work pattern will have an impact on the delivery of medical and surgical services. Quantifying this effect is necessary to plan on how to deal with COVID-19 sequelae in the recovery phase.
- **What does this study add?** As ischaemic heart disease continues to be the leading cause of death in the world, assessing the direct and indirect effect of COVID-19 on cardiology through COVID-19 related cardiac diseases and through the restriction of cardiology provisions is necessary to plan our services in the post COVID-19 era.
- **How might this impact on clinical practice?** The impact of COVID-19 will be felt beyond the direct effect of COVID-19 related cardiac diseases. The provisions of cardiac services were severely restricted due to a shift in the focus in dealing with the surge of patients with COVID-19 and patients’ reluctance to seek medical help during the lockdown period. There was therefore a reduction across all cardiology performance indicators from referrals to investigations, diagnoses and management of cardiology patients. It is therefore expected that there will be another surge of patients seeking cardiology care and that services need to plan to treat these patients early and urgently to prevent any long term complications.

## Introduction

The emergence of COVID-19 pandemic has changed the delivery of medical care across the world. In the UK, there are 280 thousand confirmed cases with more than 40000 COVID-19 related deaths.^1^ This unprecedented surge in COVID-19 infections led many governments around the world to implement population lifestyle changes through social distancing, shielding and lockdown measures to limit the spread of infection. In the UK, healthcare services had to be restructured to cope with the increased pressure and demand on the NHS.^2^

Hospitals themselves can be a potential source of infection; indeed in Lombardy hospitals became the new epicentre of infections. ^3^ Therefore, the healthcare system had to be restructured to minimise patient contact with healthcare professionals, limit or reschedule hospital visits and outpatient clinics, postpone non-urgent procedures and adopt telephone and video-assisted consultations.^4^ The UK government used an educational campaign to reduce pressure on NHS resources by advising people to ‘Stay Home; Save Lives; Protect the NHS’. In addition more extreme lifestyle changes through shielding were implemented for those deemed to be most vulnerable, such as those with cardiac transplants, complex congenital heart disease and advanced heart failure in order to reduce their exposure risk.^5^

In line with government guidelines, cardiology services had to alter the delivery of care by adopting virtual clinic models, redeployment of staff to the acute medical services and rescheduling of non-urgent procedures while at the same time dealing with the cardiac complications of COVID-19 such as myocarditis, myocardial infarction and heart failure.^6^

Across Europe the surge in COVID 19 infection is now declining giving hope that we are in the recovery phase. However, the risk of a second wave is still present. Since ischaemic heart disease is the leading cause of death, ^7^ it is necessary to assess the impact of the lockdown and healthcare restructuring on the performance of cardiology service provision and the changes needed to prepare for the recovery phase and a potential rebound surge of clinical activity.

## Methods

We set out to assess key performance indicators of cardiology services in a district general hospital (Dumfries and Galloway Royal Infirmary serving a population of 149,000) and supporting tertiary services (Golden Jubilee National Hospital, Glasgow) in the periods prior to and during the COVID-19 lockdown. We compared four time intervals, each one month long: 21 January to 20 February (baseline); 21 February to 20 March (transition period), when concerns/expectations about COVID-19 increased and service restructuring preparations were developed; 21 March to 20 April, and 21 April to 20 May (lockdown periods). Changes including reductions in cardiology service provisions and clinical activity during the lockdown intervals were compared to the first interval of baseline activity.

### Patient and Public Involvement

There has been Patient and Public Involvement (PPI) during the development of the study design and interpretation.

We consulted several service users in primary and secondary care settings through cardiac rehabilitation, heart failure, outpatient and inpatient cardiology services. They were all strongly supportive of the research.

We will continue to involve patients and the public in implementing the outcomes of this study.

## Results and Discussions

### I. Cardiology admissions and myocardial infarction

During the first month of lockdown there was a marked reduction in chest pain/breathlessness presentations with a 53% reduction in cardiology ward and Coronary Care Unit (CCU) admissions. Similarly, there was a 40% drop in the number of patients diagnosed with myocardial infarction (Table 1). Other markers were the significant fall in the number of acute cardiac tests performed, with a 46% reduction in Cardiac Troponin T (cTnT) blood tests and an 87% reduction in 12 lead ECGs. Moreover, there was a 44% reduction in inpatient echocardiograms, and 75% fewer NT pro-BNP blood tests performed both in primary and secondary care indicating a reduction in patients presenting with symptoms and signs of heart failure (Table 2).

**Table 1:**
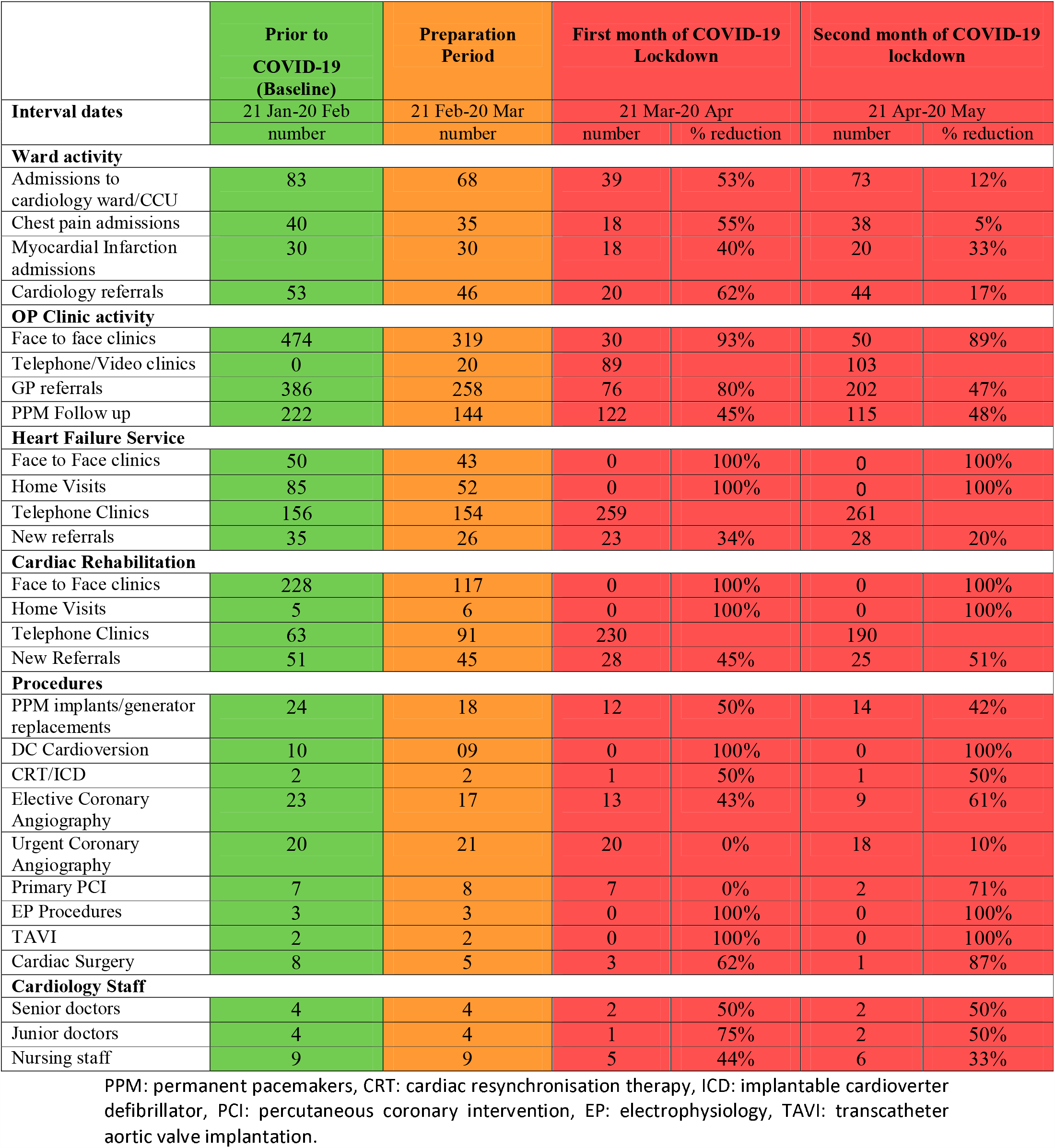
Patient reviews (inpatient and outpatient activity), procedures performed, and staff variations for different intervals: Baseline activity (green), Preparation period (amber), and during the COVID-19 lockdown. % reduction refers to the change in data from the baseline period to the period of lockdown.

**Table 2:**
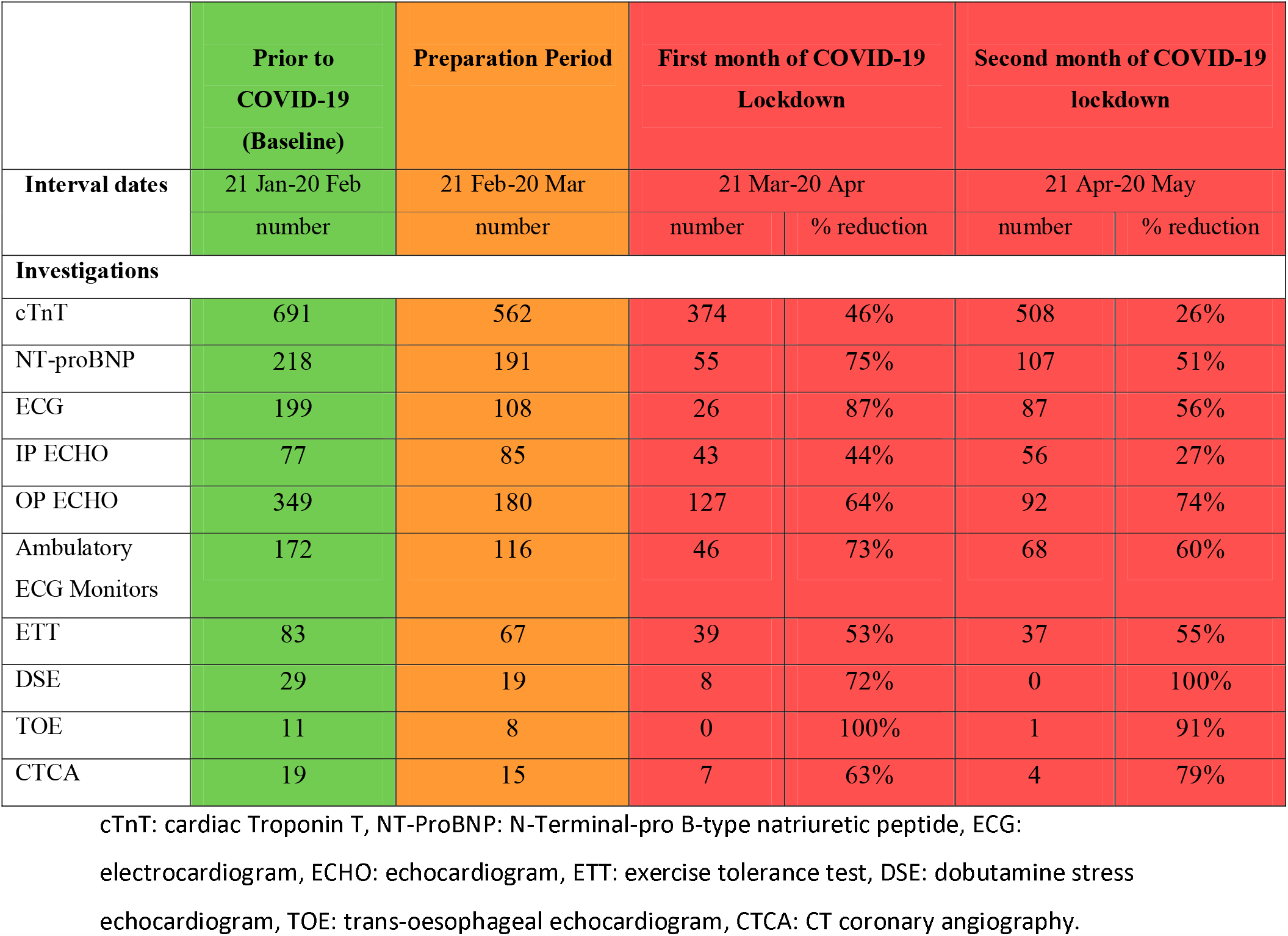
Diagnostic Activity (inpatient and outpatient): Baseline activity (green), Preparation period (amber), and during the COVID-19 lockdown. % reduction refers to the change in data from the baseline period to the periods of lockdown.

|  | Prior to COVID-19 (Baseline) | Preparation Period | First month of COVID-19 Lockdown |  | Second month of COVID-19 lockdown |  |
| --- | --- | --- | --- | --- | --- | --- |
| Interval dates | 21 Jan-20 Feb | 21 Feb-20 Mar | 21 Mar-20 Apr |  | 21 Apr-20 May |  |
|  | number | number | number | % reduction | number | % reduction |
| **Investigations** |  |  |  |  |  |  |
| cTnT | 691 | 562 | 374 | 46% | 508 | 26% |
| NT-proBNP | 218 | 191 | 55 | 75% | 107 | 51% |
| ECG | 199 | 108 | 26 | 87% | 87 | 56% |
| IP ECHO | 77 | 85 | 43 | 44% | 56 | 27% |
| OP ECHO | 349 | 180 | 127 | 64% | 92 | 74% |
| Ambulatory ECG Monitors | 172 | 116 | 46 | 73% | 68 | 60% |
| ETT | 83 | 67 | 39 | 53% | 37 | 55% |
| DSE | 29 | 19 | 8 | 72% | 0 | 100% |
| TOE | 11 | 8 | 0 | 100% | 1 | 91% |
| CTCA | 19 | 15 | 7 | 63% | 4 | 79% |

cTnT: cardiac Troponin T, NT-ProBNP: N-Terminal-pro B-type natriuretic peptide, ECG:
electrocardiogram, ECHO: echocardiogram, ETT: exercise tolerance test, DSE: dobutamine stress
echocardiogram, TOE: trans-oesophageal echocardiogram, CTCA: CT coronary angiography.

Even though these falls improved during the second month of lockdown, they were still below the baseline level of activity indicating an ongoing fall in the overall number of patients presenting to the cardiology services.

### II. Cardiology outpatient service

In line with NHS guidance on implementing COVID-19 work patterns, we adopted telephone and “Attend Anywhere” video consultation services to minimise exposure risks to patients and staff. We were able to set up secure remote working access and establish virtual clinics using Technology Enabled Care Services (TECS). New and return clinic appointments were triaged into virtual or Face to Face clinics depending on the need and urgency of the referrals. Overall, the number of patients referred from primary care to cardiology outpatient clinics dropped by 80 %. As a result of that, face to face clinics dropped by 93% and there was a substantial increase in the use of the virtual clinics (Figure 1).

**Figure 1.**
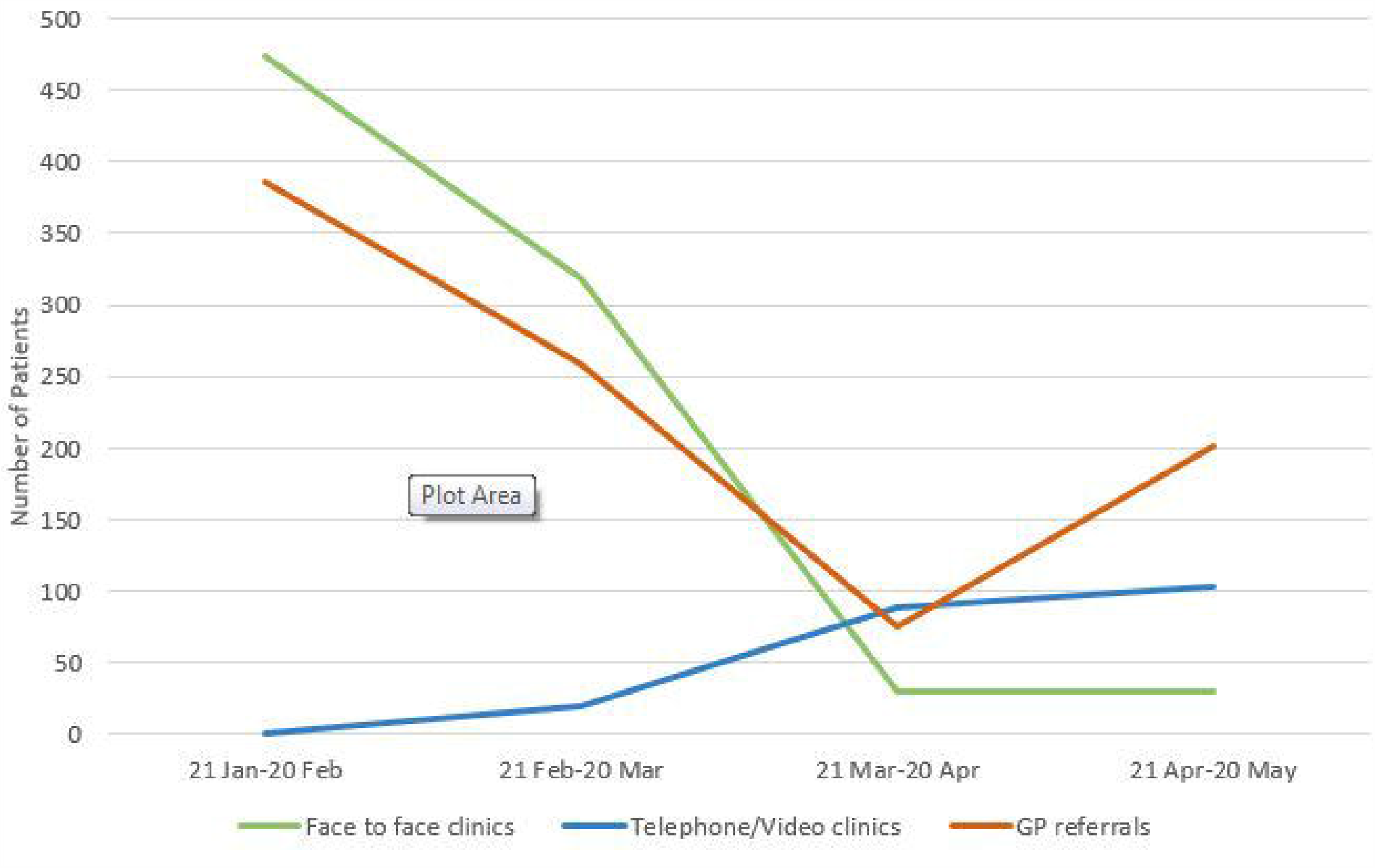
Outpatient activity before and during the lockdown intervals.

Moreover, the overriding importance of minimising potential patient exposure led to more thorough triaging of even that reduced number of primary care referrals, focussing on urgency, appropriate investigation instead of clinic assessment and telephone consultation if necessary within 24hr of receipt of the referral. In this way the majority of urgent referrals did not require face to face clinic assessment. While labour intensive at this point of entry into secondary care, this has to be a template for future outpatient practice which will yield dividends in reduced clinic waiting times once the ‘new normal’ becomes established.

### III. Cardiac Investigations

At the start of the lockdown, there was an increase in the number of patient cancellations to Clinical Physiology, and reduced face to face clinics. Many clinical physiology staff were redeployed to the critical care unit, the cardiology ward and acute medical wards to support the existing nursing staff. For instance, the hospital telemetry system became supported by the cardiac physiologist, allowing critical care nurses to focus on looking after COVID-19 patients. Within the Clinical Physiology Department social distancing was implemented in waiting areas and all staff wore droplet precautions PPE with every patient. Requests for cardiac investigations during the lockdown intervals were triaged into urgent and routine, with routine investigations rescheduled. Therefore, the number of ECGs and outpatients echocardiograms dropped by more than a half and the uptake of ambulatory cardiac monitors dropped by 73% (Table 2).

There was a parallel reduction in functional assessments of myocardial ischaemia (Exercise Tolerance Tests (ETT) and Dobutamine Stress Echocardiograms (DSE)); all routine DSEs were postponed till after lockdown. Other investigations such as Trans-Oesophageal echocardiography (TOE) and CT Coronary Angiography (CTCA) were significantly reduced, by 63% for CTCA, and cancellation of routine TOEs in line with the British Society of Echocardiography COVID-19 guidance (Figure 2). ^8^

**Figure 2.**
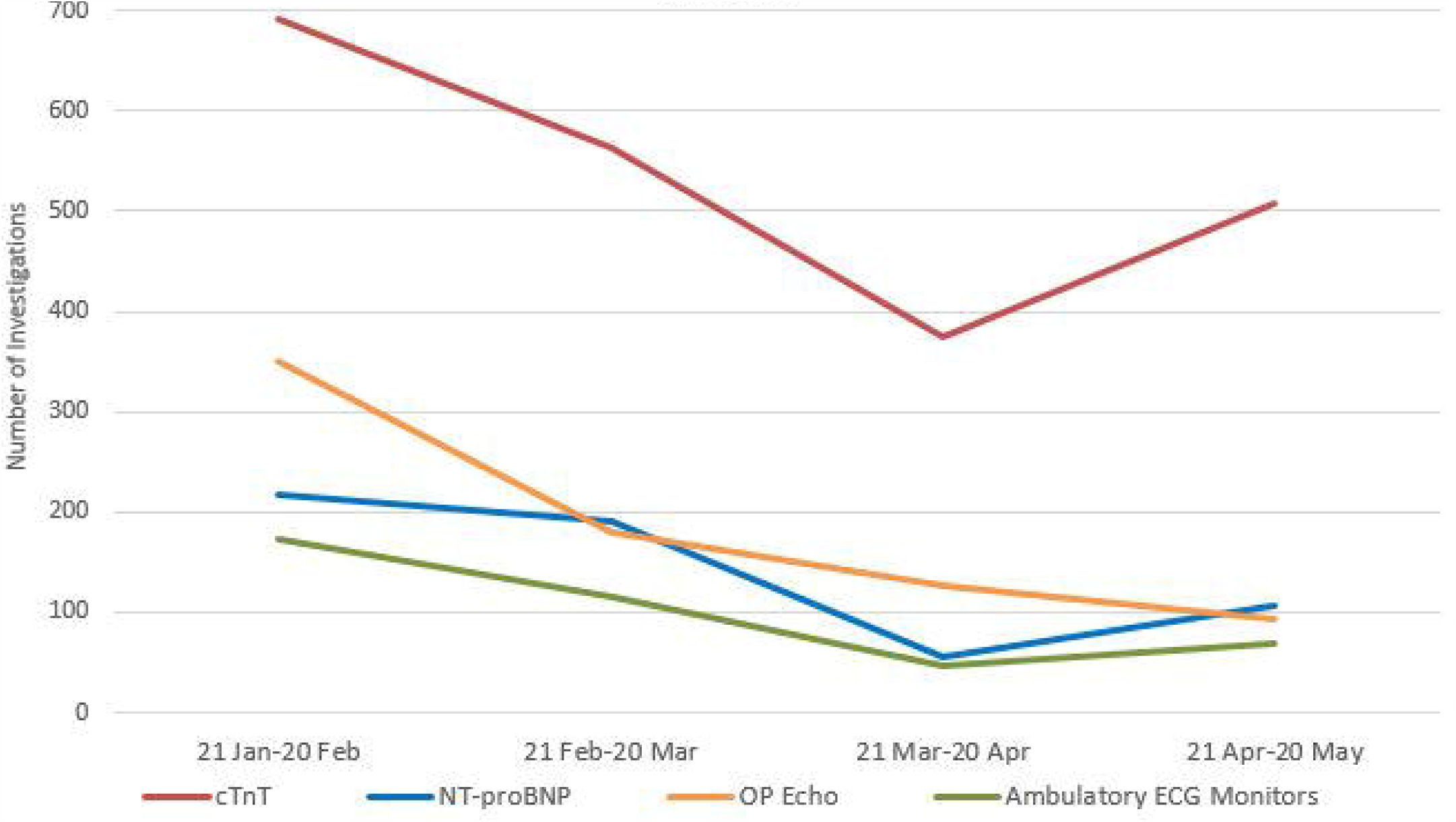
Cardiac investigations before and during the lockdown intervals.

### IV. Cardiac Procedures and Interventions

#### Cardiac Devices Implantation and DC Cardioversion

Cardiology procedures were triaged into priority groups. We used the British Heart Rhythm Society (BHRS) guidance to classify cardiac devices implantation into ‘priority 1’, ‘priority 2’ and ‘postpone’ groups.^9^ During lockdown we only performed pacemaker implantations within the priority 1 group (complete heart block or symptomatic second degree heart block). Priority 2 procedures were defined as elective procedures that could only be done during the COVID 19 period if capacity allowed, considering the risks and benefits to the patient and staff involved. These included other symptomatic bradycardia patients such as those with sinoatrial node disease or pacemaker generator replacements. Some priority 2 procedures were done during the last few weeks of the second interval of lockdown when COVID-19 new cases started to fall following consultation with patients and management. The remaining indications for pacemaker implantation were classified as ‘postpone’ and will be booked after the COVID-19 crisis is over. Therefore local device procedures halved while implantation of internal loop recorders and DC cardioversions were postponed (Table 1). Similarly, the number of patients transferred from tertiary care following complex device (CRT/ICD) implantation and electrophysiology studies dropped significantly.

Pacemaker follow up service was also reconfigured. Pacemaker patients were established on home monitoring systems and reviews were done via telephone consultation. Only patients with arrhythmias or battery depletion were brought into face to face pacing clinic for reviews.

#### Coronary Angiography and Cardiac Surgery

There was a significant drop in the number of referrals to the tertiary centre for coronary angiography and cardiac surgery. This fall was partly due to the reduction in patients presenting with chest pain and myocardial infarction but also due to the prioritisation of patients needing urgent coronary angiography. Therefore, despite a significant reduction in elective coronary angiography, there was no change in the number of urgent coronary angiography or primary PCI performed (Figure 3). A possible explanation is that patients were presenting late to hospital with more severe ischaemia requiring urgent angiography, having resisted earlier appropriate admission because of fears of contracting the virus.

**Figure 3.**
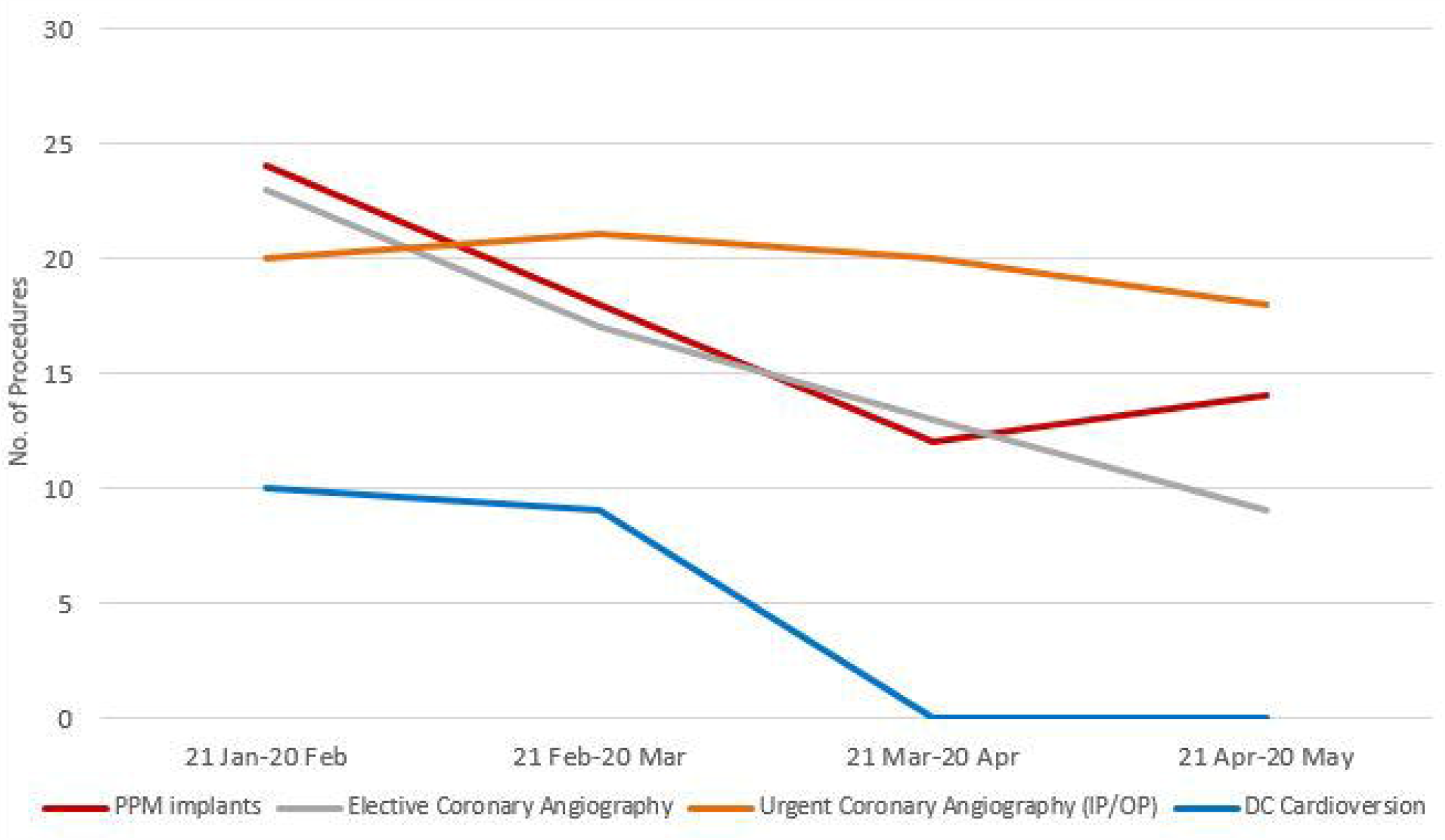
Cardiac procedures before and during the lockdown intervals.

### V. Heart Failure

COVID-19 work pattern implemented restrictions on NHS services including the community Heart Failure Service (HFS) with a cessation of face to face clinics and home visits. The Heart failure specialist nurses were maintained within the HFS. They, however, had to participate in familiarisation training with the ward environments in anticipation for potential redeployment.

New patients referred to HFS dropped by 34 % in keeping with the trends in heart failure symptom presentation, and NT pro-BNP measurements across primary and secondary care.

Home visits and face to face clinic appointments were put on hold and there was therefore an increased reliance on virtual clinics with a 66 % increase in the telephone clinics and “Attend Anywhere” video consultation services (Table 1).

In order to optimise patients’ care during the lockdown intervals, strategies to educate and empower patients were adopted such as providing patients with blood pressure monitors and scales to facilitate the uptitration and optimisation of treatment and forging even closer cooperation with primary care staff to monitor renal function.

### VI. Cardiac Rehabilitation

The cardiac rehabilitation delivery changed from the usual face to face service to a virtual service via telephone call and NHS “Attend Anywhere” video consultation. The last face to face assessments and supervised exercise sessions took place on the 16^th^ of March. The capacity of the service diminished substantially with redeployment of cardiology specialist and rehabilitation nurses to help with the reconfiguration of the cardiology ward to create a new 4 bedded Coronary Care/ High Dependency Unit thereby freeing up capacity within the critical care unit. There was therefore a 45% reduction in the delivery of cardiac rehabilitation (Table 1). There was a substantial growth in the exploration and adoption of healthcare technologies through the use of a Digital Heart Manual, the use of My Heart App and online exercise programs and the issuing of blood pressure and heart rate monitoring machines to patients through the Integrated Community Equipment Services (ICES). These new tools have successfully been used during the COVID-19 lockdown intervals and will likely continue to be a permanent part of cardiac rehabilitation after the service is restored to full capacity.

## Conclusion

To our knowledge, this is the first study in the UK to assess the impact of COVID-19 on the provision of cardiac services and clinical activity. The analysis of this UK single centre experience showed that COVID-19 pandemic has led to a significant reduction in all parts of the cardiology services including referrals to outpatient clinics, investigations, cardiology admissions, number of patients diagnosed with myocardial infarction, cardiac procedures, interventions and community services such as heart failure and cardiac rehabilitation. The findings are consistent with the recently published results of the haematology team in Oxford University Hospitals indicating a significant drop in the bloods tests performed, referrals to haematology services and diagnoses of haematological malignancies as a result of the COVID-19 lockdown measures. ^10^ Therefore, it is likely that similar changes will be seen in other medical and surgical specialties and that assessment, preparation and restructuring of these services for the recovery phase should be based on these findings.

The reasons for these falls are multifactorial and include the restructuring and prioritisation of the NHS services, reduced access to primary care and patients’ reluctance to seek medical help due to fear of contracting the virus. At the height of the pandemic, it is acceptable to deviate from the standard level of care and agreed guidelines in order to prioritise the delivery of essential services. However, adverse consequences on some patients presenting with worsening of their underlying cardiac conditions have been inevitable. Therefore, the cardiology services should be ready to offer them urgent input and early intervention.

The second interval of lockdown shows a gradual increase in patients referred to cardiology services and a rise in the investigation and diagnosis of myocardial infarction compared to the first interval of lockdown. There is therefore an expectation of a rebound surge of increased workload on cardiology services, which need to restructure in preparation for that.

The COVID-19 work pattern established technology at the heart of the delivery of care where virtual clinics using telephone and video consultation will be a long lasting legacy and MDTs and meetings using video conferencing will be the norm. It also highlighted the need to develop a communication infrastructure to make sure that these services can be delivered to all parts of the community regardless of the geographic or economic background.

The COVID-19 pandemic has also highlighted the versatility of the NHS and the readiness of its staff to adapt quickly and face new challenges. The pandemic has ignited the need to develop new pathways of care models and protocols and has started a new era of medical-nursing collaboration and resilience. The experience gained from the current outbreak will be vital in dealing with any future challenges.

## Data Availability

data available from cardiology department, Dumfries and Galloway Royal infirmary

## Acknowledgements

We would like to acknowledge Kim Heathcote and Adam Khengui for the collection of biochemical data, Cynthia Hart, Pam Houston, Linda Houston and Louise Plummer for cardiology admissions data, Fiona Ross, Mieke McKend and Jill Pagan for cardiac physiology service data and Alex McGuire and Martin Dawes for coronary angiography data (GJNH).

## Footnotes

### Contributors

The study was conceived and designed by OF, SB, RN, KM, CB and AM. Data collection was carried out by RN, CB, BD, VG and VW. Data analysis was carried out by OF, SB and AJ. The initial manuscript was compiled by OF, SB, RN, KM and CB. AM contributed substantially to critical revision of the manuscript.

### Patient consent for publication

Not required.

### Ethics approval

NHS Dumfries and Galloway Clinical Audit Approval Committee.

### Data sharing statement

No additional data are available.

## References

1. Number of coronavirus (COVID-19) cases and risk in the UK. Department of Health and Social Care, 2020. Available: https://www.gov.uk/guidance/coronavirus-covid-19-information-for-the-public [Accessed 05 June 2020].

2. NHS England. Operating framework for urgent and planned services in hospital settings during covid-19, 2020. Available: https://www.england.nhs.uk/coronavirus/wp-content/uploads/sites/52/2020/05/Operating-framework-for-urgent-and-planned-services-within-hospitals.pdf [Accessed 04 June 2020].

3. Remuzzi A, Remuzzi G. COVID-19 and Italy: what next?. Lancet 2020 ;395(10231) :1225□1228. doi:10.1016/S0140-6736(20)30627-9.

4. Coronavirus (COVID-19): nursing and community health staff guidance, Scottish Government, 2020. Available: https://www.gov.scot/publications/coronavirus-covid-19-nursing-and-community-health-staff-guidance/ [Accessed 06 June 2020].

5. PHE. Guidance on shielding and protecting people who are clinically extremely vulnerable from COVID-19. Public Health England. 2020.

6. Clerkin KJ, Fried JA, Raikhelkar J, et al. Coronavirus disease 2019 (COVID-19) and cardiovascular disease. Circulation 2020;141(20):1648–55.

7. Lozano R, Naghavi M, Foreman K, et al. Global and regional mortality from 235 causes of death for 20 age groups in 1990 and 2010: a systematic analysis for the Global Burden of Disease Study 2010. Lancet 2012;380(9859):2095–2128.

8. Clinical guidance regarding provision of echocardiography during the COVID-19 pandemic. British Society of Echocardiography, 2020. Available: https://bsecho.org/covid19 [Accessed 04 June 2020].

9. Guidance for listing of emergency and urgent elective admissions (including inter-hospital transfers), 2020. British Heart rhythm Society. Available: https://bhrs.com/wp-content/uploads/2020/03/Prioritising-cases_final.pdf [Accessed 05 June 2020].

10. Willan J, King AJ, Djebbari F, Turner GDH, Royston DJ, Pavord S, et al. Assessing the Impact of Lockdown: Fresh Challenges for the Care of Haematology Patients in the COVID-19 Pandemic. Br J Haematol 2020. doi.org/10.1111/bjh.16782.

